# Optimizing COVID-19 testing strategies on college campuses: evaluation of the health and economic costs

**DOI:** 10.1101/2022.12.04.22283074

**Authors:** Kaitlyn E. Johnson, Remy Pasco, Spencer Woody, Michael Lachmann, Maureen Johnson-Leon, Darlene Bhavnani, Jessica Klima, A. David Paltiel, Spencer J. Fox, Lauren Ancel Meyers

**Author notes:** Corresponding author: Lauren Ancel Meyers, The University of Texas at Austin, Department of Integrative Biology, 1 University Station C0930, Austin, TX 78712, USA.

## Abstract

Colleges and universities in the US struggled to provide safe in-person education throughout the COVID-19 pandemic. Testing coupled with isolation is a nimble intervention strategy that can be tailored to mitigate health and economic costs, as the virus and our arsenal of medical countermeasures continue to evolve. We developed a decision-support tool to aid in the design of university-based testing strategies using a mathematical model of SARS-CoV-2 transmission. Applying this framework to a large public university reopening in the fall of 2021 with a 60% student vaccination rate, we find that the optimal strategy, in terms of health and economic costs, is twice weekly antigen testing of all students. This strategy provides a 95% guarantee that, throughout the fall semester, case counts would not exceed the CDC’s original high transmission threshold of 100 cases per 100k persons over 7 days. As the virus and our medical armament continue to evolve, testing will remain a flexible tool for managing risks and keeping campuses open. We have implemented this model as an <u>online tool</u> to facilitate the design of testing strategies that adjust for COVID-19 conditions, university-specific parameters, and institutional goals.

**Author Summary:** As a part of the COVID-19 response team at a large public university in the US, we performed an analysis that considered together, the potential health and economic costs of different testing policies for the student body. University administrators had to weigh the up-front effort needed to implement wide scale testing against the potential costs of responding to high levels of disease on campus in the Fall of 2021, after vaccines were widely available but vaccination rates among college students were uncertain. The results presented here are applied to this specific instance, but the <u>online tool</u> provided can be tailored to university specific parameters, the epidemiological conditions, and the goals of the university. As we confront newly emerging variants of COVID-19 or novel pathogens, consideration of both the health and economic costs of proactive testing may serve as a politically tractable and cost-effective disease mitigation strategy.

## Introduction

During the first two years of the COVID-19 pandemic, universities throughout the US struggled to provide in-person education while mitigating the health and economic risks of COVID-19. The 2020-2021 academic year was particularly challenging, with many universities severely restricting in-person activities (1,2). Although the roll-out of vaccines to college-aged students in 2021 (3) ultimately allowed universities to restore many of the key elements of the residential campus experience, many spent the summer of 2021 planning for an uncertain future, as new variants emerged and vaccine uptake slowed.

Initial data on vaccine effectiveness indicated that vaccines available in the US significantly reduced the incidence of symptomatic disease (4), susceptibility to infection (5), and transmissibility if rare breakthrough infections did occur (6). Under this scenario, universities with high levels of vaccine coverage could tentatively relax face mask requirements and other precautionary measures (3,7). However, vaccine efficacy rapidly dropped with immunological waning and the emergence of new variants (8–10). Universities without vaccine mandates had to rely on estimates from vaccination data and surveys, for example in May of 2021, national polls suggested that 49% of 18-24 year olds had been vaccinated or planned to get vaccinated, but uptake varied considerably throughout the country (11), with 29% of college-aged students expressing strong hesitancy (12). Thus, many universities looked to face masks and proactive testing as low cost strategies for managing risks while reopening campus.

While college students, especially those vaccinated, are at a low risk of severe health outcomes, transmission may spillover into the surrounding community leading to surges in cases, hospitalizations and deaths. While we do not explicitly model such indirect effects, mitigating risks to vulnerable populations remains a motivating factor for preventing viral transmission on college campuses.

Several universities deployed large-scale proactive testing programs to monitor and mitigate SARS-CoV-2 activity during the 2020-2021 academic year (13–15).

Retrospective analysis suggests that these programs reduced transmission at universities (16,17) and in the surrounding communities (18). With the increasing availability and decreasing costs of SARS-CoV-2 tests, large-scale proactive testing leading to early detection and isolation of infections has become a viable but underutilized strategy for mitigating surges (17,19,20).

Here, we introduce a framework for designing cost effective testing strategies on a college campus that consider the transmission dynamics of a well-mixed, partially-vaccinated student population following a particular testing policy. A positive test drives students into isolation, where they are unable to transmit to others. The overall effectiveness of the testing policy depends on the vaccination rate, immunity from prior infection, transmissibility of the virus, vaccine effectiveness, and compliance with testing and isolation. The economic factors considered include the cost of both the proactive testing and the response and mitigation required for each positive case. These factors include the cost of a confirmatory PCR test, isolation facilities, sequencing, contacttracing, and the cost incurred to the university of needing to move classes online.

Building on prior cost effectiveness analyses of COVID-19 screening policies (21–23) and university COVID-19 policies (7,13,24), we developed this approach to support planning efforts at one of the largest public universities in the US during the summer of 2021. Using the University of Texas at Austin as a case study, we derive cost-effective testing strategies to prevent campus closures in a partially-vaccinated community of 50,000 students during the emergence of a novel variant (Delta). The model is available as an <u>online tool</u> to support universities throughout the US in tailoring COVID-19 screening programs as novel variants continue to drive waves of infection.

## Results

In the summer of 2021, we derived an optimal proactive testing strategy for the University of Texas at Austin, an urban public university with 50,000 students, for the upcoming fall semester. Given the uncertainty in vaccination rates that some universities faced, we considered a range of vaccination rates (Figure 1 and Table 1). If 60% of students arrive vaccinated, we project that cases could far surpass the CDC’s threshold for high COVID-19 activity, potentially triggering a campus closure. With passive testing (only symptom-based care seeking), we estimate that symptomatic case counts would peak between 550 and 830 (median: 700) in mid-October. If 75% of all students test two times per week the expected peak reduces to 40-100 (median: 70), with a 95% guarantee of remaining below the closure threshold. This optimal strategy would require approximately 75,000 tests per week. If 90% of students are vaccinated, however, weekly testing would be sufficient to prevent an overwhelming surge.

**Figure 1.**
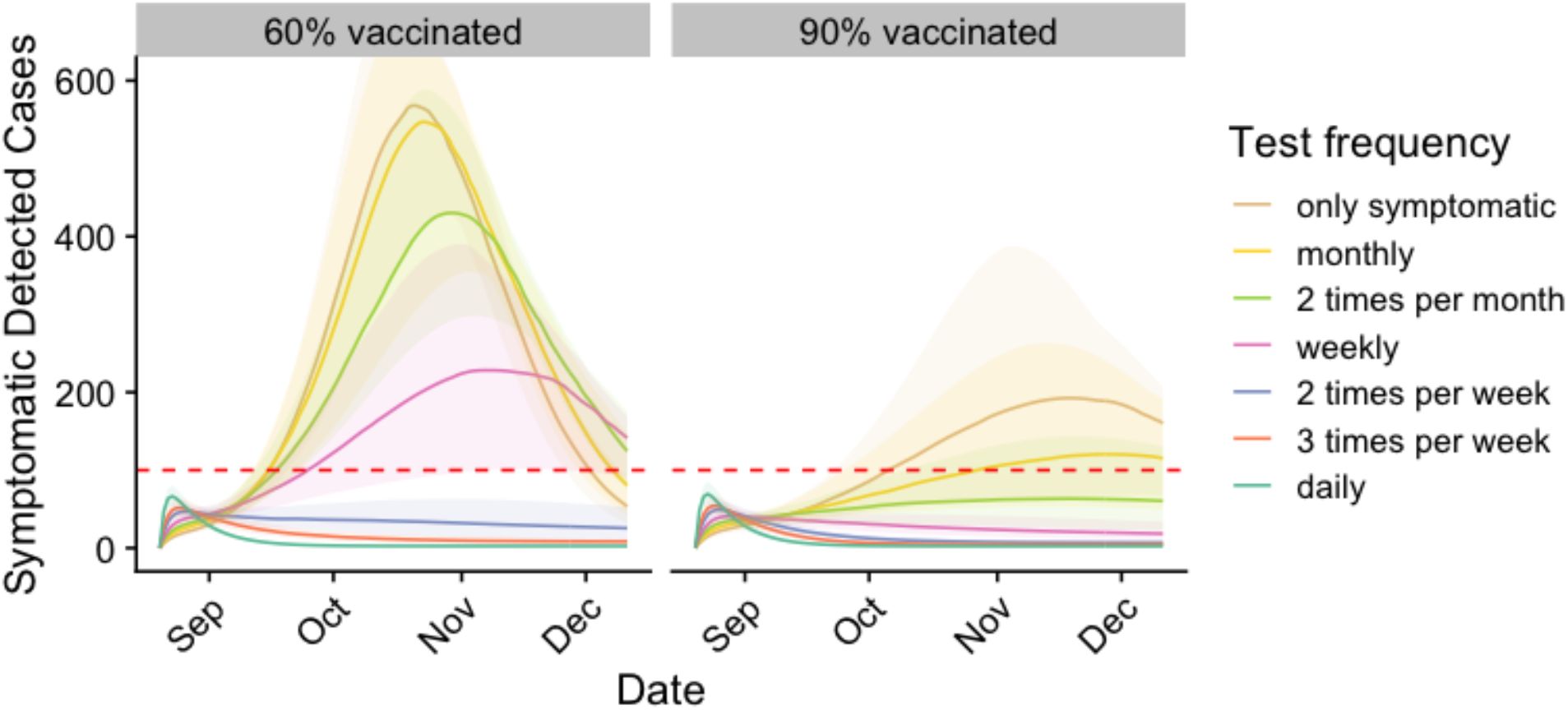
Projected COVID-19 cases among students under different levels of proactive testing, assuming 60% (left) or 90% (right) of students are fully vaccinated. Graphs project the seven-day total of detected symptomatic cases. Colors indicate testing frequency assuming 75% compliance. Shading indicates 90% prediction intervals. Horizontal lines represent the assumed campus closure threshold (twice the CDC’s high transmission threshold).

**Table 1.** Recommended testing levels under three different policy options. Testing recommendations are based on the minimum amount of testing needed to provide 95% certainty that symptomatic infections will not exceed the campus closure threshold across a range of vaccination rates.

| Population of students tested | Percent of students fully vaccinated |  |  |  |  |
| --- | --- | --- | --- | --- | --- |
|  | 50% | 60% | 70% | 80% | 90% |
| Number of tests per week |  |  |  |  |  |
| All | 3 | 2 | 2 | 2 | 1 |
| Unvaccinated at twice the rate of vaccinated | 3 | 3 | 2 | 2 | 2 |
| Only unvaccinated | 7 | daily | daily | daily | Not possible* |
| Total number of tests per week |  |  |  |  |  |
| All | 112,500 | 75,000 | 75,000 | 75,000 | 37,500 |
| Unvaccinated at twice the rate of vaccinated | 84,375 | 78,750 | 48,750 | 45,000 | 41,250 |
| Only unvaccinated | 13,1250 | 105,000 | 78,750 | 52,500 | Not possible* |

\* Due to the very small size of the population being tested, if vaccination rates are very high and testing only is occurring in the unvaccinated, it is not possible to ensure that the campus closure threshold won't be exceeded

The optimal testing frequency depends on both vaccine coverage and whether vaccinated students are exempt from testing (Table 1, Figure S4). At 90% vaccine coverage, testing only the unvaccinated would be insufficient, given our assumption that vaccines reduce risks of infection by only 47%. Across vaccination rates, exempting vaccinated students from testing requires frequent (daily) testing of unvaccinated students to prevent a surge, costing more testing resources than if all students regardless of vaccination status were tested. Testing vaccinated students at half the rate as unvaccinated students remains a viable option, as total testing resources are, at most vaccination rates, lower than if all students were tested. At 70% vaccine coverage, testing the unvaccinated 2 times per week and the vaccinated weekly requires 48,750 tests per week compared to 75,000 if vaccinated and unvaccinated test at equal rates. Across testing frequencies, the costs and infections associated with either prevention (proactive tests) or outbreak response (contact-tracing, isolation, sequencing, confirmatory PCR) are expected to be significantly higher under 60% vaccine coverage than 90% vaccine coverage (Figure 2, Figure S3).

**Figure 2.**
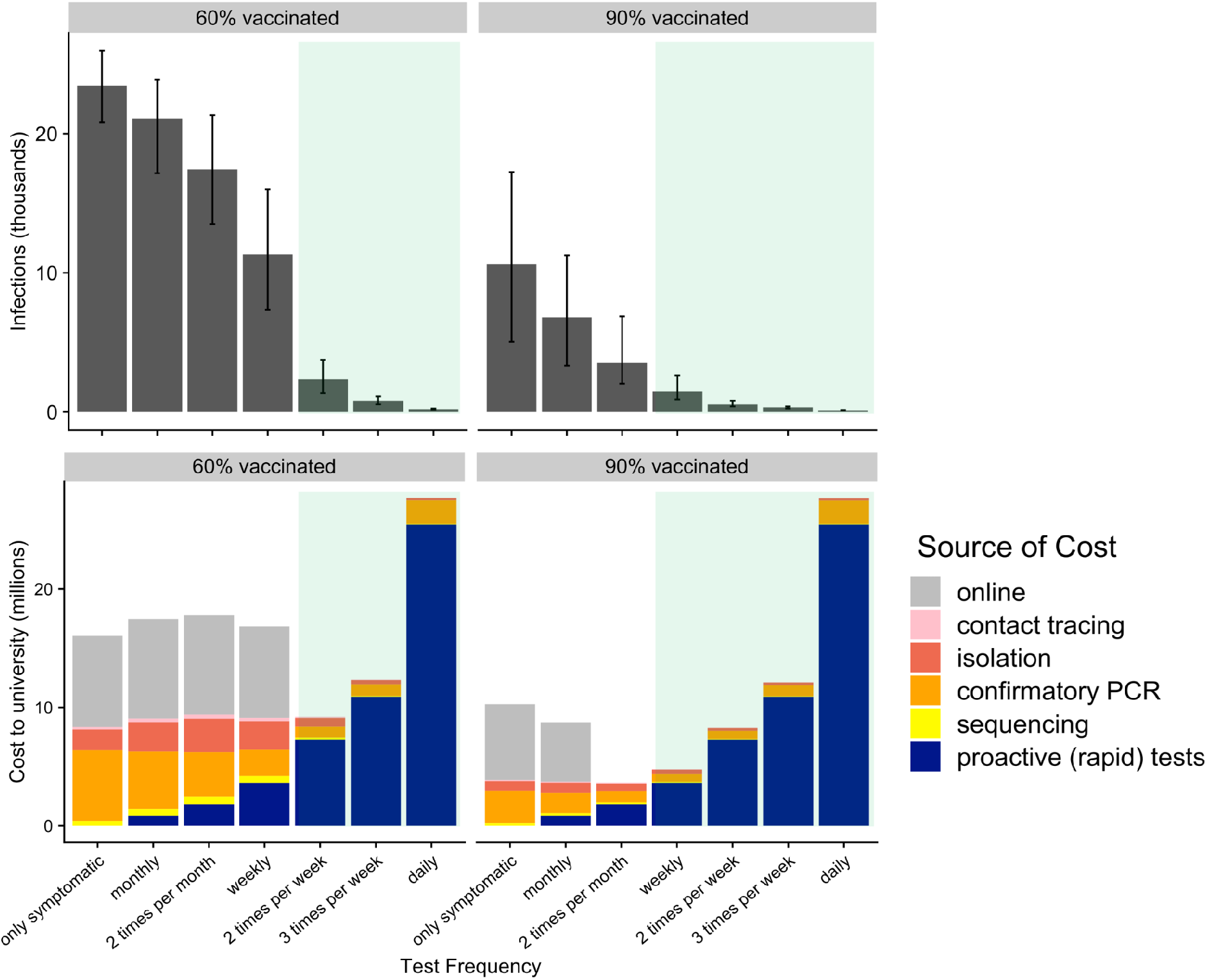
Projected health and economic costs over one semester under different levels of proactive testing, assuming 60% (left) or 90% (right) of students are fully vaccinated. Upper graphs indicate the median and 90% predictive interval of projected cumulative infections. Lower graphs indicate the projected costs, broken down by the source (colors). The green shading indicates testing frequencies that ensure the university would not exceed its closure threshold.

At 60% vaccine coverage, proactive testing of all students two times per week is sufficient to avoid exceeding the campus closure threshold at an estimated cost of around $9.1 million (Table 2). At 90% vaccine coverage, proactive testing of all students weekly is sufficient to avoid closure at a cost of $4.7 million (Table 2). We note that it costs nearly twice as much ($9.1 million vs $4.7 million, Figure 2, Table 2) to avoid campus closure at 60% vaccine coverage than at 90% vaccine coverage.

**Table 2.** Estimated level of proactive testing required to provide a 95% guarantee that detected symptomatic cases will remain below the campus closure threshold. The total cost includes the cost of the minimum proactive testing needed to stay under the threshold, plus the cost of pandemic related expenses (i.e., confirmatory testing, isolation, contact-tracing, sequencing).

|  | Percent of students fully vaccinated |  |  |  |  |
| --- | --- | --- | --- | --- | --- |
|  | 50% | 60% | 70% | 80% | 90% |
| **Minimum frequency (in all students)** | 3 times per week | 2 times per week | 2 times per week | 2 times per week | weekly |
| **Total proactive tests per week** | 112,500 | 75,000 | 75,000 | 75,000 | 37,500 |
| **Total cost to university (\$ if testing implemented)** | \$12.5 million | \$9.1 million | \$8.7 million | \$8.4 million | \$4.7 million |
| **Cost of testing per student** | \$218 | \$145 | \$145 | \$145 | \$73 |
| **Number of infections expected if only symptomatic testing is** | 25,600 | 23,600 | 20,400 | 17,000 | 10,800 |
| offered |  |  |  |  |  |

At both the 60% and 90% vaccination rates, providing the optimal amount of testing would be cost saving or nearly equivalent to the cost of the resources needed for outbreak response. At insufficient testing levels, the cost of outbreak response (i.e., contact-tracing, isolation, confirmatory PCR, and sequencing) match if not exceed the cost of proactive testing. If we assume that crossing the campus closure threshold triggers a costly move to online instruction, then proactive testing at the necessary levels is always cost-saving. When considering health costs, failing to provide sufficient testing results in significantly higher infection rates, for example, at 60% vaccination sufficient testing of 2 times per week results in 2,280 infections compared to 23,700 infections if only symptomatic testing is offered (Table 2).

## Discussion

At large US universities, large-scale proactive testing can help to suppress transmission and be cost saving overall. Although the upfront costs of proactive testing and personnel may be large, it may ultimately avert the higher costs of outbreak response and campus closure. In the fall 2021 scenarios analyzed, the costs of an effective proactive testing program per student per semester are relatively low, ranging from $73-218$ per student, depending on the vaccination rate. As we confront newly emerging variants of COVID-19 or novel pathogens, proactive testing may serve as a politically tractable and cost-effective mitigation strategy in college communities with low levels of population immunity.

Our projections suggest that, even at high vaccination rates, testing only unvaccinated students is insufficient to avoid a surge. The two other policies considered––testing all students equally and testing unvaccinated students at twice the rate of vaccinated students––are expected to exact similar overall costs and require comparable testing resources.

Large-scale asymptomatic testing is a nimble mitigation tool for universities facing novel variants, changing levels of immunity, and shifting attitudes towards face masks and intrusive social distancing measures. Testing levels can be tuned to match the changing risks and achieve university goals. When community-wide immunity is high and transmission is low, universities can reduce testing levels. As immune-evasive variants emerge and immunity wanes, universities can scale up testing to safeguard in-person activities.

Although the quantitative results of this study pertain to a specific university in the fall of 2021, the qualitative findings and modeling framework can broadly inform COVID-19 planning at US colleges and universities. We acknowledge the limitation that the cost parameters used in this analysis are based on the situation at this specific university, rather than from nationally representative cost estimates. Because these costs, as well as the epidemiological and university specific parameters, will vary widely over time and space, we have developed an interactive online tool (28) to facilitate generalizability of the analysis. The spread and costs of COVID-19 will depend not only on the factors considered here, but also on university policies, student behavior, vaccine uptake throughout the semester, and the emergence of variants with different levels of transmission, immune evasiveness, and severity.

We note that our projections assume a high and constant transmission rate throughout the simulation period, and thus do not account for changes in face mask usage, social distancing, and contact tracing efforts. The model assumes that 75% of students would participate in proactive testing, which would require aggressive health communication and outreach. The model also assumes that individuals fully isolate following a positive test, which may require the provision of additional isolation rooms, paid sick-leave, and removal of academic penalties for missed classes. Additionally, the model does not explicitly consider the effects of waning immunity (from vaccination or prior infections), though we address this in the shiny app by allowing the user to input the percent of students optimally immunized (i.e. up to date on their booster shots). We have not considered the health or economic costs of severe disease, long COVID, or death within the university community nor how these risks may differ between students, faculty, and staff, nor do we consider the cost to the individual of missing class due to isolation resulting from a positive test.

Prior studies have demonstrated that frequent rapid testing can reduce transmission (7,13,17,20) and is cost-effective (21–23). A similar decision-support tool helps universities optimize testing while keeping cumulative cases below 5% of the population (7). The CDC (29) and ACHA (3) have continually released guidance to help universities navigate the rapidly changing COVID-19 situation. Building on these contributions, our study offers a tool that incorporates the health effects, and also the economic effects, not just of the cost of the testing program but also the costs of responding to positive COVID-19 cases in a university setting.

## Materials & Methods

### Transmission model

We analyzed a compartmental model of SARS-CoV-2 transmission that incorporates vaccination and isolation following a positive test. A full description of the model structure and parameters are provided in the Supplement (Section S1). In our case study, we modeled COVID-19 during the summer of 2021, immediately after the Delta variant rose to dominance. We assumed that vaccines reduce the risks of infection by 47% [95% CI: 37-50%] (8,25), reduce the likelihood of developing symptoms by 64% [95% CI: 63-73%] (25), and reduce transmission to others by 20% (10). We assumed a reproduction number (*R*_0_) of 5, without interventions, that 75% of students comply with testing policies, and that students who test positive isolate for 7 days. We did not explicitly model the effect of quarantining close contacts on reducing transmission. Finally, we assumed that 25% of symptomatic individuals infected with SARS-COV-2 would seek testing. We tracked the rolling seven-day total detected symptomatic cases, where cases are detected through both proactive (antigen) testing and symptom-based care seeking.

### Economic model

To estimate the costs of testing, we considered both testing supplies and the personnel needed to administer tests, collect data, and process results. All proactive testing was assumed to be performed via antigen testing, at a significantly reduced cost than PCR tests. We assumed that all positive proactive tests received a PCR confirmatory test.Symptom-based care seekers received only a PCR test. PCR confirmation was then followed by contact tracing, molecular sequencing of the test specimen, and a seven-day isolation period. At the time of the case study, contact-tracing was being performed to encourage students who were close contacts of positive cases to get tested. We assumed that 20% of positive cases require a campus-provided isolation room, based on internal data from the university. Finally, we considered the costs of campus closures triggered by large surges. Based on conversations with university leadership, we assumed that on-line instruction incurs additional costs totalling $100,000 per day. We do not explicitly consider educational losses (i.e. missed class) or administrative costs of coordinating COVID-19 responses, nor do we directly consider the healthcare costs associated with student illness

### Campus closure thresholds

We assumed that universities would revert to hybrid or online instruction when case counts surpassed the following public health thresholds (26).

- High risk: 100 detected symptomatic cases per 100,000 people in a seven-day period, corresponding to the original CDC red (high) alert level.
- Higher risk: 150 detected symptomatic cases per 100,000 people in a seven-day period, corresponding to the 1.5 times the original CDC red (high) alert level.
- Very high risk: 200 symptomatic detected cases per 100,000 people in a seven-day period, corresponding to double the original CDC red (high) alert level.

Our case study assumed that the university would close when the seven-day new symptomatic case count exceeded the very high risk threshold. In our online tool, we provide even higher thresholds to support universities in mitigating highly transmissible variants with lower severity, like Omicron (27).

### Identification of optimal testing levels

We considered a range of vaccination rates, from 50% to 90% vaccinated in 10% increments. For a given level of vaccination, we identified the minimum amount of proactive testing required to ensure that the university does not exceed its closure threshold, with a 95% guarantee. For each candidate policy, we ran 100 deterministic simulations, each with parameters randomly selected from their specified distributions (Table S2), and identified the policy with the least amount of testing in which 95% of simulations remain under the closure threshold. We note that we identified the optimal policy conditioned on the vaccination rate; across vaccination rates, the costs associated with either proactive testing or outbreak response generally increase as the vaccination rate decreases.

### Sensitivity analyses

Beyond vaccination rates, testing frequencies, and risk tolerances, several other factors influence the projections. To elucidate their impact, we conducted a sensitivity analysis with respect to vaccine effectiveness against infection (ranging from 40%-90%, base case at 47%), vaccine effectiveness against onwards transmission if infected (ranging from 50% to 0% effective, base case at 20%), and the testing policy (only unvaccinated, unvaccinated at double the rate of vaccinated, and all students equally). The results are provided in the Supplement (Figure S5).

## Data Availability

All code and data used to make the manuscript figures are available at this Github repo: https://github.com/kej1993johnson/campus_testing_cost_effectiveness

## Data availability statement

All code and data used to make the manuscript figures are available at this <u>Github repo</u>. A separate <u>Github repo</u> is available here to deploy the latest version of the Rshiny app.

## Acknowledgments

We acknowledge the financial support from NIH Grant R01 AI151176, CDC Grant U01IP001136, and funding from Tito’s Handmade Vodka. All authors declare no conflicts of interest. We would like to thank all of the individuals involved in the Proactive Community Testing team at the University of Texas at Austin.

## Supporting Information Captions

S1 Text. COVID-19 Transmission model with vaccination and testing

S1 Figure. Compartmental model of COVID-19 transmission incorporating testing and vaccination.

S1 Table. Initial conditions

S2 Table. Transmission model parameters

S3 Table. Cost parameters

S2 Text. Results for vaccination coverage ranging from 50% to 90% with all students tested

S2 Figure. Projected COVID-19 cases among students under different levels of proactive testing, assuming 50%, 60%, 70%, 80%, and 90% vaccination coverage amongst students.

S3 Figure. Projected health and economic costs through December 16, 2021 under different levels of proactive testing, assuming 50%, 60%, 70%, 80% or 90% of students are fully vaccinated

S3 Text. Results for vaccine coverage ranging from 50% to 90% and different populations tested

S4 Figure. Projected COVID-19 cases among students under different levels of proactive testing, assuming 50%, 60%, 70%, 80%, and 90% vaccination coverage and in testing policies for all students, testing vaccinated at half the rate, and testing in the unvaccinated only.

S4 Table. Estimated level of proactive testing to provide 95% guarantee that symptomatic cases will remain below the *very high risk threshold* if testing at half the rate in vaccinated vs unvaccinated.

S5 Table. Estimated level of proactive testing to provide 95% guarantee that symptomatic cases will remain below the *very high risk threshold* if only the unvaccinated are tested.

S3 Text. Sensitivity analysis: vaccine efficacy against infection and transmission S5 Figure. Projected COVID-19 cases among students as a function of the vaccine efficacy against infection and symptomatic disease assuming 50%, 60%, 70% or 80% of students are fully vaccinated.

S6 Figure. Projected COVID-19 cases among students as a function of the vaccine efficacy against transmission assuming 50%, 60%, 70% or 80% of students are fully vaccinated.

S4 Text. Modifications to framework/Rshiny app for future variants

S6 Table. Rshiny app default settings and suggested adjustments for Omicron.

