## Supplement for "Optimizing COVID-19 testing strategies on college campuses: evaluation of the health and economic costs"

#### S1 Text. COVID-19 transmission model with vaccination and testing

The model structure is diagrammed in Figure S1 and described in the equations below. In short, the population is divided into four groups based on vaccination status (subscripts $v$ and $u$) and quarantine status (subscript $q$). Within these groups, individuals can transition between disease states: susceptible ($S$), exposed ($E$), infectious pre/asymptomatic ($I_a$), infectious symptomatic ($I_s$), and recovered ($R$). The symbols $S$, $E$, $I_a$, $I_s$, and $R$ denote the number of people in that state in the given vaccination/quarantine group. Individuals can transition from the active (in contact with others) state to the quarantine state and back based on receiving a positive test result and being released from quarantine, respectively. The total size of the active population (anyone not in quarantine) at any time is given by:

$$X(t) = S_u + E_u + I_{au} + I_{su} + R_u + S_v + E_v + I_{av} + I_{sv} + R_v.$$

The force of infection at time t, denoted as $\beta^*(t)$ is given by:

$$\beta^*(t) = \beta_A(I_{au} + e_v I_{av}) + \beta_S(I_{su} + e_v I_{sv})$$

Where $\beta_A$ and $\beta_S$ are the transmission rate of asymptomatic and symptomatic individuals respectively, and $e_v$ is the reduction in transmissibility of vaccinated infected individuals.

The model equations governing transition from one state to the next are given by:

$$\frac{dS_u}{dt} = -\beta^*(t)\frac{S_u}{X(t)} - S_u w_u(1 - Sp)if_u + S_{qu}k - in$$

$$\frac{dE_u}{dt} = \beta^*(t)\frac{S_u}{X(t)} - \gamma E_u - E_u w_u(1 - Sp)if_u$$

$$\frac{dI_{au}}{dt} = \gamma E_u - \delta_{au} I_{au} - I_{au} w_u Seif_u + E_{qu} k + in$$

$$\frac{dI_{su}}{dt} = p_{sym}\delta_{au} I_{au} - \delta_s I_{su} - i_{sym} I_{su} - I_{su} w_u Seif_u$$

$$\frac{dR_u}{dt} = \delta_s I_{su} + (1 - p_{sym})\delta_{au} I_{au} - R_u w_u(1 - Sp)if_u + R_{qu}k + \delta_q I_{squ} + \delta_q I_{aqu}$$

$$\frac{dS_{qu}}{dt} = S_u w_u(1 - Sp)if_u - S_{qu}k$$

$$\frac{dE_{qu}}{dt} = E_u w_u(1 - Sp)if_u - E_{qu}k$$

$$\frac{dI_{aqu}}{dt} = I_{au} w_u Seif_u - \delta_q I_{aqu}$$

$$\frac{dI_{squ}}{dt} = I_{su} w_u Seif_u + i_{sym} I_{su} - \delta_q I_{squ}$$

$$\frac{dR_{qu}}{dt} = R_u w_u(1 - Sp)if_u - R_{qu}k$$

$$\frac{dS_v}{dt} = -\sigma_v \beta^*(t)\frac{S_v}{X(t)} - S_v w_v(1 - Sp)if_v + S_{qv}k$$

$$\frac{dE_v}{dt} = \beta^*(t)\frac{S_v}{X(t)} - \gamma E_v - E_v w_v(1 - Sp)if_v$$

$$\frac{dI_{av}}{dt} = \gamma E_v - \delta_{av} I_{av} - I_{av} w_v Seif_v + E_{qv}k$$

$$\frac{dI_{sv}}{dt} = p_{sym,v}\delta_{av} I_{av} - \delta_s I_{sv} - i_{sym} I_{sv} - I_{sv} w_v Seif_v$$

$$\frac{dR_v}{dt} = \delta_s I_{sv} + (1 - p_{sym,v})\delta_{av} I_{av} - R_v w_v(1 - Sp)if_v + R_{qv}k + \delta_q I_{sqv} + \delta_q * I_{aqv}$$

$$\frac{dS_{qv}}{dt} = S_v w_v(1 - Sp)if_v - S_{qv}k$$

$$\frac{dE_{qv}}{dt} = E_v w_v(1 - Sp)if_v - E_{qv}k$$

$$\frac{dI_{aqv}}{dt} = I_{av}w_vSeif_v - \delta_q I_{aqv}$$

$$\frac{dI_{squ}}{dt} = I_{sv}w_vSeif_v + i_{sym}I_{sv} - \delta_q I_{sqv}$$

$$\frac{dR_{qv}}{dt} = R_v w_v(1 - Sp)if_v - R_{qv}k$$

Where $\sigma_v$ is the relative susceptibility to infection if vaccinated, $Se$ and $Sp$ are the sensitivity and specificity of surveillance tests, $w_v$ and $w_u$ are the willingness to test amongst the vaccinated and unvaccinated individuals respectively, $f_v$ and $f_u$ are the surveillance testing frequencies among vaccinated and unvaccinated individuals respectively, $i$ is the probability that an individual isolates after receiving a positive test result, $i_sym$ is the probability that an individual isolates and seeks testing after developing symptoms, $k$ is the rate of confirmatory testing, $\gamma$ is the transition rate from exposed to infectious calculated from the latent period duration, $p_{sym}$ and $p_{sym,v}$ are the proportion of infected individuals who eventually show symptoms in the unvaccinated and vaccinated groups respectively, $\delta_{au}$ and $\delta_{av}$ are the transition rates out of the asymptomatic compartment for the vaccinated and unvaccinated individuals, respectively, and $\delta_s$ and $\delta_{sq}$ are the transition rates out of the symptomatic compartments for the active and quarantined individuals respectively. The values of the transition rates are estimated from the average time spent in each compartment. See Table S2 for all parameter values. The model is implemented as a system of differential equations that are solved in R using the deSolve package [1]. It is assumed that the entire population of active individuals, including vaccinated and unvaccinated individuals are well mixed. The initial conditions are provided in Table S1, the values for the parameters in the transmission model are provided in Table S2, and the cost assumptions are provided in Table S3. Uncertainty was incorporated into the model via parameter uncertainty only. In short, we ran 100 simulations for each scenario we were interested in. At each simulation, we sampled from the parameter distributions prescribed, and ran the deterministic model forward. Summary statistics were calculated based on the results either at each time point or as an endpoint from eac simulation.

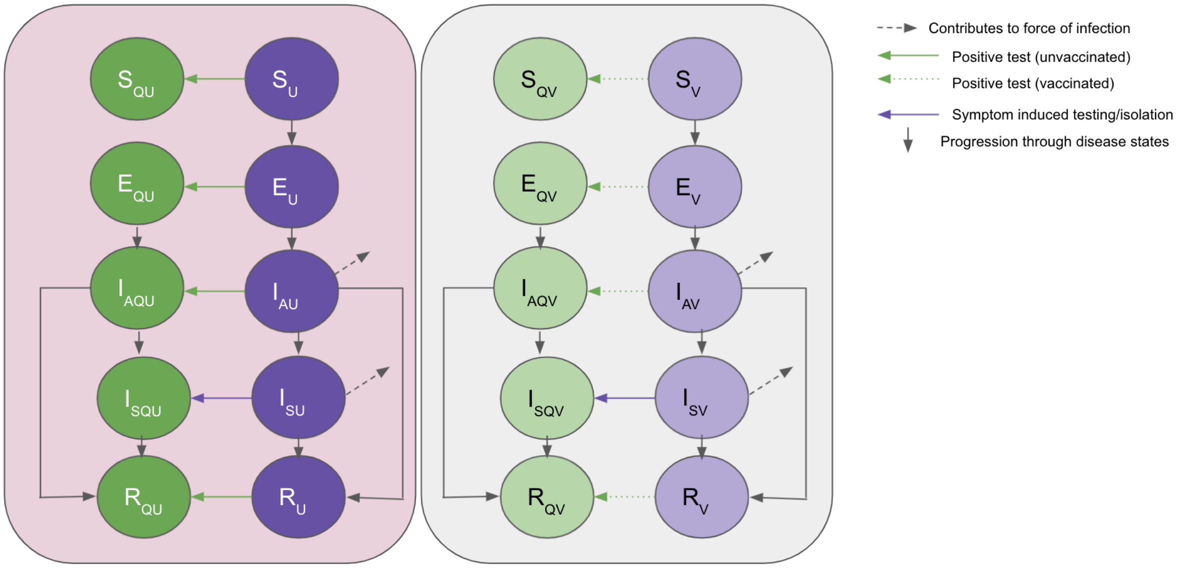


**S1 Figure. Compartmental model of COVID-19 transmission incorporating testing and vaccination.** Each group (defined by vaccination ($v$, $u$) and quarantine states ($q$)) is modeled with a set of compartments. Upon infection, susceptible individuals ($S$) progress to the exposed compartment ($E$) and then to asymptomatic infectious compartment ($I_a$). Some of those progress to symptomatic infections ($I_s$) and some go directly to recovered ($R$). Testing frequencies dictate the rate at which individuals move into their corresponding disease states in the quarantine group.

S1 Table. Initial conditions

| **Variable** | **Settings** |
|---|---|
| Initial infection prevalence among UT students | 420 [380-470] per 100,000 based on introduction estimates for the University of Texas at Austin [2] |
| Initial fraction immune among students | Triangular (32%, 40%, 48%) based on CDC seroprevalence estimates for TX [3] + hospitalizations from June 15-Aug 03, 2021 |
| Number of students | 50,000 |

S2 Table. Transmission model parameters

| **Parameter** | **Value** | **Source** |
|---|---|---|

| | | |
|---|---|---|
| $e_v$: Relative transmissibility of infected individuals that have been vaccinated | 0.8 | [4] |
| $\sigma_v$: relative susceptibility to infection if vaccinated | Triangular(0.37, 0.47, 0.5) corresponding to (63%, 53%, 50%) efficacy at preventing infection | [5,6] |
| $sym_{red,v}$: overall reduction in chance of symptomatic disease if vaccinated | Triangular (27%, 36%, 37%) corresponding to (73%, 64%, 63%) effective at preventing symptomatic disease | [6] |
| $S_p$: specificity of antigen test | 99.5% | test specificity [7–9] |
| $S_e$: sensitivity of antigen test | 90% for antigen | test sensitivity [7–9] |
| $w_v$: test acceptance rate in vaccinated individuals | 75% | Assumed |
| $w_u$: test acceptance rate in unvaccinated individuals | 75% | Assumed |
| $f_v$: daily frequency of test offer to vaccinated individuals | Varied from once every month to daily | Assumed |
| $f_u$: daily frequency of test offer to unvaccinated individuals | Varied from once every month to daily | Assumed |
| *i:* isolation probability for individuals who receive a positive test result or who develop symptoms | 92.5% | Assumed |
| $i_{sym}$: isolation probability for individuals who develop symptoms | Triangular(24%, 25%, 38%) | Assumed |

| | | |
|---|---|---|
| *k:* rate of confirmation testing | 1/2 | Assumed based on time to seek test after rapid result + PCR test-turnaround time |
| $t_{exposed}$: *duration of latent period* | 3 days | [10] |
| $t_{presym}$: duration of pre-symptomatic period | 2.3 days | [10] |
| $t_{infectious}$: total duration of infectiousness (same for both asymptomatic and presymptomatic) | 7 days | [10,11] |
| $t_{symptomatic}$: *duration of symptomatic infectiousness* | 4.7 days | $t_{infectious}$ - $t_{presym}$ |
| $t_{quarantine}$: *duration of quarantine for positive individuals* | 7 days | [12] |
| $p_{sym}$: proportion of infectious individuals that eventually show symptoms | Triangular(0.5, 0.6, 0.7) | [13] |
| $p_{sym,v}$: proportion of vaccinated infectious individuals that eventually show symptoms | Triangular(0.54, 0.76, 1 ) | Calculated based on reduction in susceptibility ($\sigma_v$) and reduction in overall symptomatic probability of those vaccinated $p_{sym,v} = \frac{sym_{red}}{\sigma_v}$ |
| $\delta_s$: recovery rate from symptomatic infection | 1/4.7= 0.21 | Calculated from the average time in the symptomatic infectious state |
| $\delta_{au}$: transition rate out of asymptomatic unvaccinated compartment | $\frac{1}{t_{asym,u}}$= Triangular(0.12, 0.14, 0.17 ) | Calculated based on the average time in asymptomatic compartment and the proportion symptomatic |

| | | |
|---|---|---|
| | | $t_{asym,u} = p_{sym}t_{presym} + (1 - p_{sym})t_{infectious}$ ,<br>which implies<br>$\delta_{au} = (t_asym, u)^{-1}$ |
| $\delta_{av}$: transition rate out of asymptomatic vaccinated compartment | $\frac{1}{t_{asym,v}}$ = Triangular(0.03, 0.04, 0.06 ) | Calculated based on the average time in asymptomatic compartment and the proportion symptomatic<br>$t_{asym,v} = p_{sym,v}t_{presym} + (1 - p_{sym,v})t_{infectious}$<br>which implies<br>$\delta_{av} = (t_asym, v)^{-1}$ |
| $\delta_q$*: rate of release from quarantine if true positive* | 1/7 = 0.14 | Calculated from time in the symptomatic quarantined state |
| $R_0$*:* basic reproductive number | Triangular(4.5, 5, 5.5) | Wildtype $R_0$ ~ 2.7 [14] Alpha 60% more transmissible than wildtype [15], Delta 60% more transmissible than Alpha [16] |
| $\beta$: daily transmission rate of infectious individuals | Triangular(0.64, 0.71, 0.79) | Calculated from $R_0$ , $\beta = \frac{R_0}{t_{infectious}}$ |
| *in:* number of introductions per week | Triangular(7.5, 10, 12.5) | Assumed |

S3 Table. Cost parameters

| Variable | Setting | Source |
|---|---|---|
| Confirmatory PCR test | $100 per test | [17,18] |
| Sequencing of positive sample | $100 per sample | [19] |
| Contact-tracing | $50 per positive | Conversations with head of contact-tracing at UT Austin (Darlene Bhavnani) |
| Isolation facility usage | $300 per student per day | Conversations with UT Austin administrators (Johnathan Robb) |
| | 20% of students testing positive use isolation facility | Based on UT percent of on campus housing who use isolation facility plus others living off campus (will be much higher if most students live on-campus) |
| | 7 day isolation period | Assumed from duration of infectiousness |
| Rapid surveillance tests | $6 including staffing | Conversations with UT Austin test coordinators |
| Cost per day of moving classes | $100,000 per day | Conversations with UT Austin administrators (Art Markman and John Salsman, who indicated moving online would require computers to students plus additional facilities and other disruptions. |

### S2 Text. Results for vaccination coverage ranging from 50% to 90% with all students tested

In this section, we provide epidemiological projections for vaccination scenarios ranging from 50% to 90% vaccination rate of the student body, with testing policies either targeted to all students equally (main text results), half as much in vaccinated students, and in unvaccinated only.

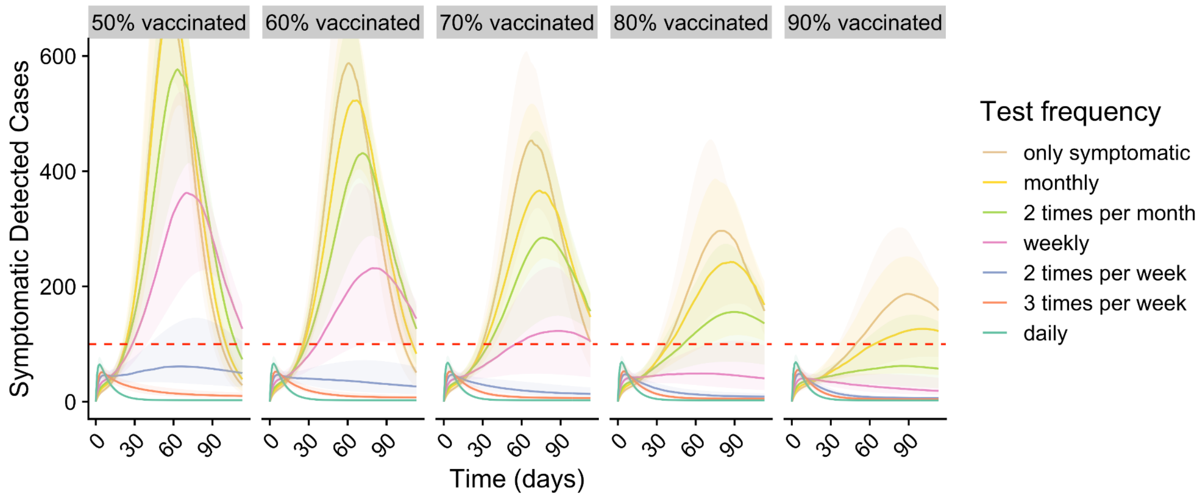


**S2 Figure. Projected COVID-19 cases among students under different levels of proactive testing, assuming 50%, 60%, 70%, 80%, and 90% vaccination coverage amongst students.** Graphs project the daily prevalence of symptomatic infections detected through a combination of symptomatic and proactive testing. Colors indicate the testing frequency for all students, assuming 75% compliance. Shading indicates the 90% prediction intervals. The red dashed horizontal line represents the *very high risk threshold*.

Focusing on the main text scenario where all students are tested at equal rates regardless of vaccination status, we show the full breakdown of cost and infection projections for vaccination rates ranging from 50% to 90%.

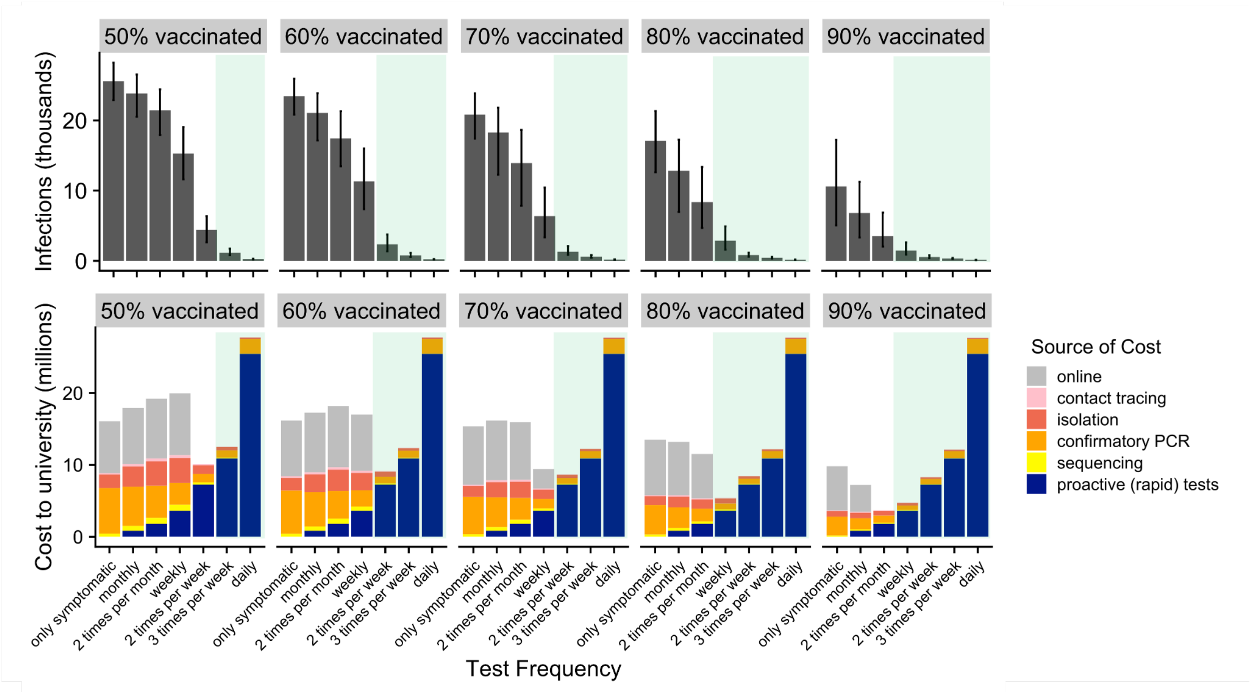


**S3 Figure. Projected health and economic costs through December 16, 2021 under different levels of proactive testing, assuming 50%, 60%, 70%, 80% or 90% of students are fully vaccinated.** The top graphs indicate the median and 90% predictive interval of projected cumulative infections.The bottom graphs indicate the projected costs, broken down by the source (colors). The green shading indicates testing frequencies that are expected to keep symptomatic prevalence below the *very high risk threshold*.

### S3 Text. Results for vaccine coverage ranging from 50% to 90% and different populations tested

The main text results focus on the testing policy in which vaccinated and unvaccinated are tested at the same rate. However, it is reasonable that decision-makers would also want to explore alternative policies, such as testing only the unvaccinated or testing vaccinated half as much as the unvaccinated. In this section, we present the results of the projected symptomatic detected cases for all three of these testing policies side by side (Figure S4).

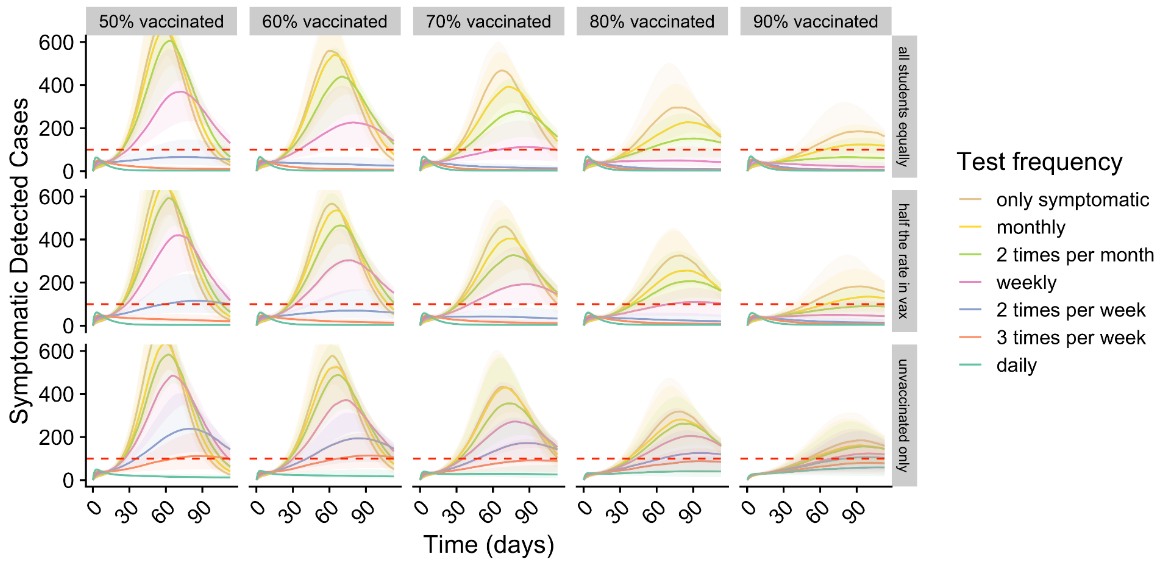


**S4 Figure. Projected COVID-19 cases among students under different levels of proactive testing, assuming 50%, 60%, 70%, 80%, and 90% vaccination coverage and in testing policies for all students, testing vaccinated at half the rate, and testing in the unvaccinated only.** Graphs project the daily prevalence of symptomatic infections detected through a combination of symptomatic and proactive testing. Colors indicate the testing frequency for all students, assuming 75% compliance. Shading indicates the 90% prediction intervals. The red dashed horizontal line represents the *very high risk threshold*. The top row corresponds to the testing of all students at an equal rate, the middle row corresponds to testing vaccinated at half the rate of unvaccinated, and the bottom row corresponds to testing of the unvaccinated only.

In the main text, we provide a table with the testing recommendation, total proactive tests, total cost to the university and cost (of testing) per infection averted assuming we test all students at the same rate. We provide the companion tables for the testing policies in which vaccinated are tested at half the rate as unvaccinated and vaccinated only are tested.

**S4 Table. Estimated level of proactive testing to provide 95% guarantee that symptomatic cases will remain below the *very high risk threshold* if testing at half the rate in vaccinated vs unvaccinated.**

| | **Percent of students fully vaccinated** | | | | |
|---|---|---|---|---|---|
| | **50%** | **60%** | **70%** | **80%** | **90%** |
| **Minimum frequency in unvaccinated** | 3 times per week | 3 times per week | 2 times per week | 2 times per week | 2 times per week |

| | | | | | |
|---|---|---|---|---|---|
| **students** | | | | | |
| **Minimum frequency in vaccinated students** | 3 times every 2 weeks | 3 times every 2 weeks | weekly | weekly | weekly |
| **Total proactive tests per week** | 84,375 | 78,750 | 48,750 | 45,00 | 41,250 |
| **Total cost to university ($)** | $9.7 million | $8.9 million | $5.9 million | $5.4 million | $4.9 million |
| **Cost (of testing) per infection averted ($)** | $345 | $343 | $260 | $290 | $400 |
| **Cost of testing per student** | $163 | $152 | $94 | $87 | $80 |
| **Number of infections expected if only symptomatic testing is offered** | 25,600 | 23,600 | 20,400 | 17,000 | 10,800 |

**S5 Table. Estimated level of proactive testing to provide 95% guarantee that symptomatic cases will remain below the *very high risk threshold* if only the unvaccinated are tested.**

| | **Percent of students fully vaccinated** | | | | |
|---|---|---|---|---|---|
| | **50%** | **60%** | **70%** | **80%** | **90%** |
| **Minimum frequency in unvaccinated students** | daily | daily | daily | daily | * |
| **Minimum frequency in vaccinated students** | never | never | never | never | * |
| **Total proactive tests per week** | 131,250 | 105,000 | 78,750 | 52,500 | * |
| **Total cost to university ($)** | $14.7 million | $12.1 million | $9.7 million | $7.1 million | * |
| **Cost (of testing) per infection averted ($)** | $526 | $476 | $421 | $376 | * |

| **Cost of testing per student** | $254 | $203 | $152 | $102 | * |
|---|---|---|---|---|---|
| **Number of infections expected if only symptomatic testing is offered** | 25,600 | 23,600 | 20,400 | 17,000 | 10,800 |

*indicates that even daily testing in the unvaccinated isn't sufficient to keep symptomatic cases below the very high risk threshold with 95% certainty. This is because at high vaccination levels, this corresponds to a very small percent of the student body.

#### S3 Text. Sensitivity analysis: vaccine efficacy against infection and transmission

In the main results, we used the most up-to-date estimates of vaccine efficacy against infection, symptomatic disease, and transmission to make the health and economic projections. However, these estimates changed significantly from the time we initiated this project to when we finished it, and we expect that estimates of vaccine effectiveness will continue to be updated with new data and potentially with the impact of new variants. To better understand how our results hinge on this uncertainty, we performed a sensitivity analysis on vaccine effectiveness against infections and symptomatic disease. For each vaccination rate ranging from 50 to 90% coverage, we assumed weekly testing was present at 75% participation in all students regardless of vaccination status. We then varied vaccine efficacy against infection from 40% to 70% in 10% increments, and vaccine efficacy against symptomatic disease from 55% to 85% also in 10% increments. For each condition, uncertainty on vaccine efficacy against infection ranged from  5% below the median value to 22% above the median value, consistent with what is present in our main results where vaccines are assumed to be 53% effective against infection at median [5], with an uncertainty range from 50% effective to 63% effective [6].

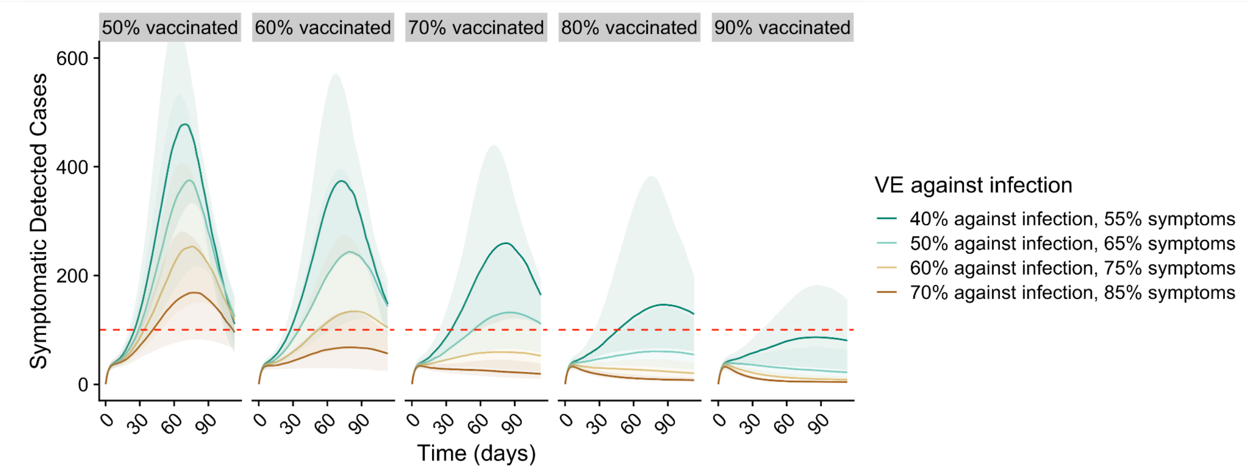


**S5 Figure. Projected COVID-19 cases among students as a function of the vaccine efficacy against infection and symptomatic disease assuming 50%, 60%, 70% or 80% of students are fully vaccinated**. Graphs project the daily prevalence of symptomatic infections detected through a combination of symptomatic and proactive testing. The projections assume weekly testing in all students as the baseline scenario. Shading indicates the 90% prediction intervals. Horizontal red dashed line represents the *very high risk threshold*.

The results of the sensitivity analysis reveal the large degree to which the results depend upon vaccine efficacy. For example, in the 70% vaccination scenario, weekly testing is more than sufficient to prevent exceeding the very high risk threshold if vaccines are 70% effective against infection and 85% effective against symptoms. However, if vaccines are only 50% effective against infection, as has recently been indicated [5], then weekly testing is projected to cause symptomatic cases to exceed the very high risk threshold. This critical uncertainty explains why analysis performed in the summer of 2021[2] presents recommendations that differ from the main results in this report. This is one of the motivating factors for releasing the Rshiny app alongside this report, allowing for the various parameters such as vaccine efficacy to be updated as the situation continues to evolve.

Another key uncertainty in vaccine efficacy is the degree to which vaccinated individuals, if infected, go on to transmit to others. Early reports indicated that breakthrough infections reduced transmissibility by 50% [20], but more recent reports have indicated that vaccinated infected individuals have similar viral loads to unvaccinated infected individuals [4]. We performed a sensitivity analysis varying the effect of vaccines on transmissibility from breakthrough infections having the same transmissibility to being 50% less transmissible.

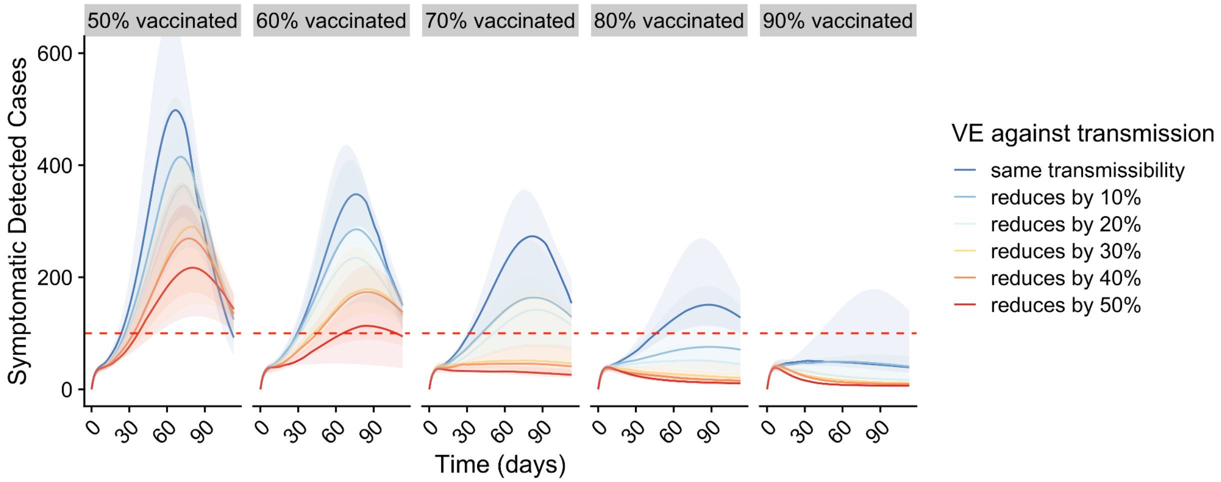


**S6 Figure. Projected COVID-19 cases among students as a function of the vaccine efficacy against transmission assuming 50%, 60%, 70% or 80% of students are fully vaccinated**. Graphs project the daily prevalence of symptomatic infections detected through a combination of symptomatic and proactive testing. The projections assume weekly testing in all students as the baseline scenario. Shading indicates the 90% prediction intervals. Horizontal red dashed line represents the *very high risk threshold*.

The results of the sensitivity analysis indicate that the degree to which vaccinated individuals transmit to others has a large effect on the resulting outbreak. For example, if vaccinated infected individuals have the exact same transmissibility as unvaccinated infected individuals, then even at 90% vaccination rate, weekly testing does not guarantee that symptomatic cases will not exceed the *very high risk threshold*. This result again underscores the need to be adaptive in testing strategies and willing to adjust input assumptions as more data about vaccine efficacy becomes available.

### S4 Text. Modifications to framework/Rshiny app for future variants

The default setting of the partner Rshiny app is parameterized to a 50,000 student university, originally intended for use for university administrators for the Fall of 2021. At this time, the Delta variant was the dominant circulating variant, and “fully vaccinated” was defined as 2 doses of the mRNA vaccines or 1 dose of an adenovirus vaccine. In order to adapt this tool for use with different variants, for example the Omicron variant, which is the dominant variant circulating at the time of submission in early 2022, we suggest the following changes:

**S6 Table. Rshiny app default settings and suggested adjustments for Omicron.**

| Parameter | Current default settings (for Delta as of Fall 2021) | How to adjust for Omicron (early 2022) |
|---|---|---|
| Percent optimally immunized | 60% using optimally immunized as having had 2 doses of an mRNA vaccine or 1 dose of an adenovirus vaccine | 20% using those having been fully vaccinated and boosted |
| Percent immune from prior infection | 32% using CDC seroprevalence surveys as of September 2021 | Because Omicron appears to evade immunity from infection from prior variants significantly, use estimates of the proportion recovered from recent infection to estimate |
| Basic reproductive number | 5 for Delta | 6.5+ for Omicron |
| Efficacy of optimal immunization against infection | 50% against any infection for Delta | Estimated around 50-60% for booster shot efficacy against Omicron infection |
| Tolerance for symptomatic cases | 2x CDC high (200 cases per 100k in 7 days) | Potential to move up to 3 or 4x the CDC high transmission level to account for potential decreased severity of Omicron |

We acknowledge that the parameter options presented to adjust in the app do not represent the complexity of all things to be considered in the face of a novel variant, for example, multiple different levels of immunity (i.e. fully vaccinated but not boosted, recovered but greater than 6 months ago, etc). Our hope is that the framework is flexible enough to allow a user to make reasoned estimates of the range of some of the parameters, and to be able to adjust them as new data emerges and specific to their university's demographics and epidemic scenario. Additionally, all code to build the app and generate all figures, along with code to estimate some campus specific parameters, is available at https://github.com/kej1993johnson/university_testing_vax_proj.git .

12. Du Z, Pandey A, Bai Y, Fitzpatrick MC, Chinazzi M, Pastore y Piontti A, et al. Comparative cost-effectiveness of SARS-CoV-2 testing strategies in the USA: a

modelling study. The Lancet Public Health. 2021;6: e184–e191.

13. Davies NG, Klepac P, Liu Y, Prem K, Jit M, CMMID COVID-19 working group, et al. Age-dependent effects in the transmission and control of COVID-19 epidemics. Nat Med. 2020;26: 1205–1211.

14. parameter_estimates/2019_novel_coronavirus at master · midas-network/COVID-19. Github; Available: https://github.com/midas-network/COVID-19

15. Davies NG, Abbott S, Barnard RC, Jarvis CI, Kucharski AJ, Munday JD, et al. Estimated transmissibility and impact of SARS-CoV-2 lineage B.1.1.7 in England. Science. 2021;372. doi:10.1126/science.abg3055

16. Allen AH, Vusirikala A, Flannagan J, Twohig KA, Zaidi A, COG-UK Consortium, et al. Increased household transmission of COVID-19 cases associated with SARS-CoV-2 Variant of Concern B.1.617.2: a national case- control study.

17. Free Coronavirus testing for privately insured patients? 20 Apr 2020 [cited 29 Sep 2021]. Available: https://www.kff.org/policy-watch/free-coronavirus-testing-for-privately-insured-patients/

18. COVID-19 test prices and payment policy - Peterson-KFF Health System Tracker. 28 Apr 2021 [cited 29 Sep 2021]. Available: https://www.healthsystemtracker.org/brief/covid-19-test-prices-and-payment-policy/

19. Simonetti M, Zhang N, Harbers L, Milia MG, Brossa S, Huong Nguyen TT, et al. COVseq is a cost-effective workflow for mass-scale SARS-CoV-2 genomic surveillance. Nat Commun. 2021;12: 3903.

20. Layan M, Gilboa M, Gonen T, Goldenfeld M, Meltzer L, Andronico A, et al. Impact of BNT162b2 vaccination and isolation on SARS-CoV-2 transmission in Israeli households: an observational study. bioRxiv. medRxiv; 2021. doi:10.1101/2021.07.12.21260377
